# Contact without protection: high antenatal attendance, minimal hepatitis B testing in Nigeria’s mother-to-child prevention cascade — how far from elimination?

**DOI:** 10.64898/2026.09.15.26363185

**Authors:** Samson Ikenna Abanni, Bifom Melwin Ekure, Jean Omotosho

## Abstract

Hepatitis B is a leading preventable cause of liver cancer, and in high-burden settings most chronic infection is acquired around birth. Nigeria carries Africa’s largest hepatitis B burden. Preventing mother-to-child transmission requires a chain of antenatal steps: testing pregnant women, acting on the result, and vaccinating the newborn. No nationally representative study has assembled these into a single prevention cascade. We built the first national hepatitis B mother-to-child prevention cascade for Nigeria and measured how far it falls from the World Health Organization’s triple-elimination targets.

We analysed the 2023-24 Nigeria Demographic and Health Survey, the first Nigerian survey to carry both a maternal hepatitis B module and infant birth-dose records. Among 13,341 women with a most recent live birth in the preceding three years, each linked to that child’s immunisation record, we estimated coverage at each cascade step (antenatal attendance, hepatitis B testing in antenatal care, result received, and the infant birth dose) using complex-survey methods, and examined patterning by wealth, education, residence, travel time, and region. Although 72% of mothers attended antenatal care, only 12.3% (95% CI 11.4-13.2) were tested for hepatitis B there, and among the few tested almost all received a result: the loss fell at testing, not at result-return. The infant birth dose reached 55.0% (53.1-56.9), 35 points below target, and ran at a level unrelated to maternal testing, consistent with universal, non-targeted delivery; place of delivery, not maternal testing, was the dominant correlate of an infant receiving it. Both steps were steeply patterned: antenatal testing ranged from 2.6% in the poorest wealth quintile to 32.4% in the richest, and was lowest in the North-West, the zone with Nigeria’s highest prevalence.

Nigeria already reaches most mothers through antenatal care but rarely uses that contact to test for hepatitis B, and the universal birth dose is only half delivered. The country is far from elimination, and furthest where the virus is commonest. Integrating hepatitis B testing into antenatal care, made free and weighted toward the highest-burden north, would convert contact already achieved into protection.

## Introduction

Hepatitis B is one of the few cancers we know how to prevent before it begins. Chronic infection with the hepatitis B virus is a principal cause of cirrhosis and hepatocellular carcinoma, and in 2018 the virus accounted for an estimated 360,000 new cancers worldwide, the third-largest share attributable to any infection, after *Helicobacter pylori* and human papillomavirus [1]. That burden is not spread evenly: sub-Saharan Africa carries among the highest rates of infection-attributable cancer of any world region, and Nigeria alone bears the largest hepatitis B burden in Africa: a national meta-analysis of 47 studies puts pooled prevalence at 9.5% [2], upwards of twenty million people [3]. What sets this cancer apart is when its story starts. In high-prevalence settings most chronic infection is established not in adulthood but in the first days and years of life, and the earlier infection takes hold the more likely it is to persist: around nine in ten babies born to mothers positive for the hepatitis B surface or e antigen develop chronic infection [4]. The liver cancer of the fifth decade is, in effect, written at the cradle, which is why interrupting transmission from mother to child is the pivot on which both hepatitis B elimination and much of Africa’s liver-cancer prevention turn.

A tool to break this chain has existed for decades. A dose of hepatitis B vaccine given within twenty-four hours of birth, followed by the routine infant series, prevents most perinatal transmission [5] and is among the most cost-effective interventions available in low-income settings [6]. Yet the birth dose is not, on its own, enough. In a Cameroonian cohort of children born to infected mothers, 5.6% were chronically infected by follow-up even when the vaccine arrived on time within twenty-four hours of birth. That residual risk fell almost entirely on one group. Among e-antigen-positive mothers with high viral load, 32.4% of children were infected despite a timely dose, against 0.3% or less in every other maternal stratum [7]. These are the mothers a vaccine cannot fully shield against, and the only way to find them is to look before delivery. Reaching the elimination target therefore turns on a sequence of steps rather than a single injection: a pregnant woman must know that hepatitis B exists, be tested for it in antenatal care, receive and understand her result, be offered antiviral prophylaxis if she carries high risk, and then see her newborn vaccinated at birth and through the infant series. Each link conditions the next, and a break at any one of them lets transmission through. It is precisely this chain that the World Health Organization’s framework for the triple elimination of mother-to-child transmission of HIV, syphilis, and hepatitis B renders as explicit coverage targets. Every country is held to at least 90% birth-dose coverage. Where the birth dose is given selectively, or where universal timely coverage has not been attained, a further target applies: at least 90% of pregnant women tested for hepatitis B surface antigen [8].

What is known about these steps is known one step at a time. National programmes report how many newborns receive the birth dose and, where it is measured, how many pregnant women are tested for hepatitis B, but as separate indicators, never joined at the level of the individual mother and child. The evidence on bringing the steps together is thinner than the consensus that they should be. It rests largely on small implementation pilots, and on reviews that synthesise programme descriptions rather than measure the population. A 2023 scoping review concludes that selective birth-dose vaccination cannot interrupt transmission without hepatitis B screening integrated into antenatal care [4]. A 2026 systematic review of integrated triple-elimination services finds the evidence fragmented and calls explicitly for evaluation of “the full cascade” from testing through infant vaccination; for West Africa, it notes, such evidence is absent [9]. The one study to link a mother’s hepatitis B status to her infant’s birth dose in Nigeria did so in a single state, among a self-selected cohort of women recruited through churches. It could speak to serological status and its immediate downstream, and to nothing else: not to whether women had heard of the virus, were tested in antenatal care, or received their results, and not to the country as a whole [3]. Missing, then, is the object that would show where the population actually falls away, and whether testing ever reaches the newborn: a nationally representative prevention cascade that follows mothers from antenatal care through hepatitis B testing and its result to their infant’s birth dose.

The 2023–24 Nigeria Demographic and Health Survey makes that object measurable for the first time. Alongside its record of childhood immunisation, the survey asked women of reproductive age whether they had heard of hepatitis B, whether and where they had been tested, and whether testing had taken place during antenatal care. This is the first time a Nigerian national survey has put those questions to mothers whose infants’ immunisation records it also holds, and so the first time a mother’s engagement with the virus can be linked to her own child’s protection at the scale of a country. We use it to build a national hepatitis B mother-to-child prevention cascade, adapting to perinatal hepatitis B the cascade-of-care framework developed to locate where patients are lost along the multi-step HIV service [10]. We trace the population from antenatal care through testing and its result to the infant’s birth dose, hold each step against the World Health Organization’s triple-elimination coverage targets [8], and locate the point at which the pathway breaks. Because Nigeria offers the birth dose universally, the question is whether the widely used antenatal platform is being used to test mothers for hepatitis B at all (the step on which any risk-targeted prevention depends), and whether a mother’s testing is met with any additional protection for her newborn beyond that universal dose. We then examine where, and for whom, the cascade breaks, by geographic access to care and region and across household wealth and education, to establish whether the distance to elimination is also a distance of equity. We find an antenatal platform widely used but rarely used for hepatitis B testing, a birth dose reaching barely half of infants, and a cascade that fails most where the virus is most common.

## Materials and Methods

### Data and design

We analysed the 2023–24 Nigeria Demographic and Health Survey (NDHS), the most recent round of the Demographic and Health Surveys programme in Nigeria, conducted between December 2023 and May 2024 by the National Population Commission and the Federal Ministry of Health with technical support from ICF. The survey used a stratified, two-stage cluster design: 1,400 enumeration areas were drawn from the national sampling frame with probability proportional to size, and households were sampled within each selected area. All women aged 15–49 in sampled households were eligible for the individual questionnaire, which completed 39,050 interviews; children born in the years preceding the survey were covered by a birth-history and immunisation module reported by their mothers. We drew on three linked sources: the women’s recode, for the maternal hepatitis B module and sociodemographic characteristics; the children’s recode, for infant vaccination; and the geospatial covariate extract, for a cluster-level measure of travel time to the nearest urban centre. The NDHS is publicly available in de-identified form and received ethical approval from Nigeria’s National Health Research Ethics Committee and the ICF Institutional Review Board; no further approval was required for this secondary analysis of public data.

### Analytic populations

Although the maternal hepatitis B module was put to all 39,050 women, its items followed the questionnaire’s skip pattern: every woman was asked whether she had heard of hepatitis B; only those who had were asked about lifetime testing and its result; and the questions on testing during antenatal care were asked of women who had given birth in the recall period and attended antenatal care. We accordingly defined two populations. Population-level awareness and lifetime testing were estimated among all women aged 15–49. The mother-to-child prevention cascade, the study’s focus, was estimated among women whose most recent live birth fell in the three years before the survey, the reference period of the childhood-immunisation module, each linked to that youngest child through the household and respondent identifiers. Because antenatal testing, the reported result, and the child’s birth dose all refer to the same pregnancy and child, their temporal order is fixed by design: antenatal testing precedes the birth it is meant to protect. Anchoring the cascade to the most recent birth aligned the mother’s antenatal report with the specific child’s vaccination record and limited recall error.

### Cascade steps and definitions

Following the cascade-of-care approach [10], we expressed the pregnancy-anchored prevention pathway as an ordered series of coverage steps, each the proportion of all recent mothers reaching that step: attendance at antenatal care (at least one visit); a hepatitis B test during that care; receipt of a definite positive or negative result; and the infant’s receipt of the birth dose, taken from the child’s vaccination card where seen and from the mother’s report otherwise (children aged 0–35 months). We report awareness of hepatitis B (ever having heard of the virus) and lifetime testing among all women as population context rather than cascade steps, since neither is a prerequisite for provider-initiated antenatal testing. “Don’t know” and missing responses were treated as the step not reached, a deliberately conservative rule that, given self-report, bounds each coverage estimate from above and the cascade’s losses from below. Self-reported antenatal positivity was rare (79 women), so the clinically important downstream step, linkage of high-risk mothers to antiviral prophylaxis, was too sparse to model and is reported descriptively rather than analysed.

### Supplementary comparison

Because Nigeria delivers the birth dose universally rather than on the basis of maternal status, we did not analyse it as an outcome of testing. As a supplementary check on whether any risk-targeted delivery was nonetheless discernible, we compared card-documented birth-dose coverage between infants of tested and untested mothers using modified Poisson regression with robust variance [11], adjusting for maternal age, education, household wealth, region, residence, and travel time. We made no causal claim, interpreting the estimate by its magnitude and precision, and computed an E-value [12] to gauge its sensitivity to unmeasured confounding.

### Geographic access

We measured geographic access with travel time to the nearest urban centre, a modelled cluster-level covariate available for every cluster in the DHS geospatial extract, analysed in quintiles. To protect confidentiality, cluster coordinates are randomly displaced before release: up to two kilometres in urban clusters and five in rural ones, with a further one per cent of rural clusters moved up to ten. Analysing access categorically limited sensitivity to this displacement. Region was the geopolitical zone and residence the urban–rural classification as coded in the survey.

### Estimation

All analyses accounted for the survey’s complex design. Estimates were weighted with the individual sampling weight for woman-level quantities and the corresponding child weight for the birth-dose and cascade analyses, with primary sampling units and stratification incorporated through Taylor-series linearisation. Cascade coverage, overall and within subgroups, was reported as a weighted proportion with a 95% confidence interval and set beside the World Health Organization’s triple-elimination targets of at least 90% antenatal testing and at least 90% birth-dose coverage. Which targets bind depends on how a country delivers the birth dose [8]. Nigerian coverage falls well short of universal and the survey does not record timeliness, placing the country in the group to which the maternal testing target applies; the cascade was therefore held to both. The cascade coverage estimates were the primary analysis; the geographic and socioeconomic patterning of cascade completion was a pre-specified secondary analysis. Analyses were conducted in R using the survey package, which implements the stratified two-stage design directly: weighted point estimates with Taylor-series linearised variance. Prevalence ratios were estimated by modified Poisson regression with a log link and design-based robust variance. The script that reproduces every value reported here is given in S5 File.

### Sensitivity and robustness

We probed the findings in two pre-specified ways. We re-estimated antenatal testing coverage treating “don’t know” responses as a separate category rather than as the step not reached, to confirm that our conservative coding did not drive the conclusion that testing falls far short of target. We also restricted the supplementary birth-dose comparison to card-documented vaccination, to gauge the influence of maternal recall.

### Ethics statement

This study is a secondary analysis of the publicly available, de-identified 2023–24 Nigeria Demographic and Health Survey. The survey received ethical approval from the National Health Research Ethics Committee of Nigeria and the ICF Institutional Review Board, and written informed consent was obtained from participants by the survey implementers. No additional ethical approval was required for this secondary analysis of public, de-identified data.

### Use of artificial intelligence

Claude (Anthropic), model claude-opus-5, was used to assist for the R analysis script and in editing the manuscript text. It was not used to generate data, results, or references, and is not listed as an author. The script in S5 File reproduces every estimate reported here and prints a verification table against each reported value; The authors verified all analyses and take full responsibility for the content.

## Results

### Analytic sample

Of the 39,050 women aged 15–49 interviewed, 13,341 had a most recent live birth in the three years before the survey and were linked to that child’s immunisation record, forming the analytic sample for the prevention cascade. Their median age was 29 years; 61% lived in rural areas and 46% had no formal education. Reflecting Nigeria’s higher fertility in the north, the sample was concentrated in the North-West (39%) and North-East (20%) and skewed toward poorer households (24% in the poorest wealth quintile against 16% in the richest; Table 1).

**Table 1.** Characteristics of the analytic sample (recent mothers with a most recent live birth in the three years before the survey), NDHS 2023–24.

| Characteristic | Weighted % (N = 13,341) |
| --- | --- |
| Median age, years | 29 |
| *Residence* |  |
| Urban | 38.7 |
| Rural | 61.3 |
| *Education* |  |
| None | 45.7 |
| Primary | 11.4 |
| Secondary | 32.5 |
| Higher | 10.4 |
| *Wealth quintile* |  |
| Poorest | 24.0 |
| Poorer | 22.2 |
| Middle | 20.0 |
| Richer | 18.0 |
| Richest | 15.7 |
| *Geopolitical zone* |  |
| North-West | 38.5 |
| North-East | 19.9 |
| North-Central | 16.7 |
| South-East | 6.7 |
| South-South | 7.8 |
| South-West | 10.4 |

### The prevention cascade

Contact with antenatal care did not translate into hepatitis B testing. Seven in ten mothers (72%) attended antenatal care at least once, yet only 12.3% (95% CI 11.4–13.2) were tested for hepatitis B during that care; among the few tested, nearly all learned their result (11.9% of all mothers), so the loss fell overwhelmingly at testing itself rather than at result-return. Set against the World Health Organization’s triple-elimination target of at least 90% antenatal testing, this is a gap of almost 78 percentage points. The infant birth dose reached 55.0% (53.1–56.9) of children, 35 points short of the 90% target, and was delivered at a far higher level than maternal testing, consistent with its universal, non-targeted administration (Fig 1). Awareness was itself limited: only 57% of these mothers had heard of hepatitis B, and across all women aged 15–49 just 6.9% had ever been tested.

**Fig 1.**
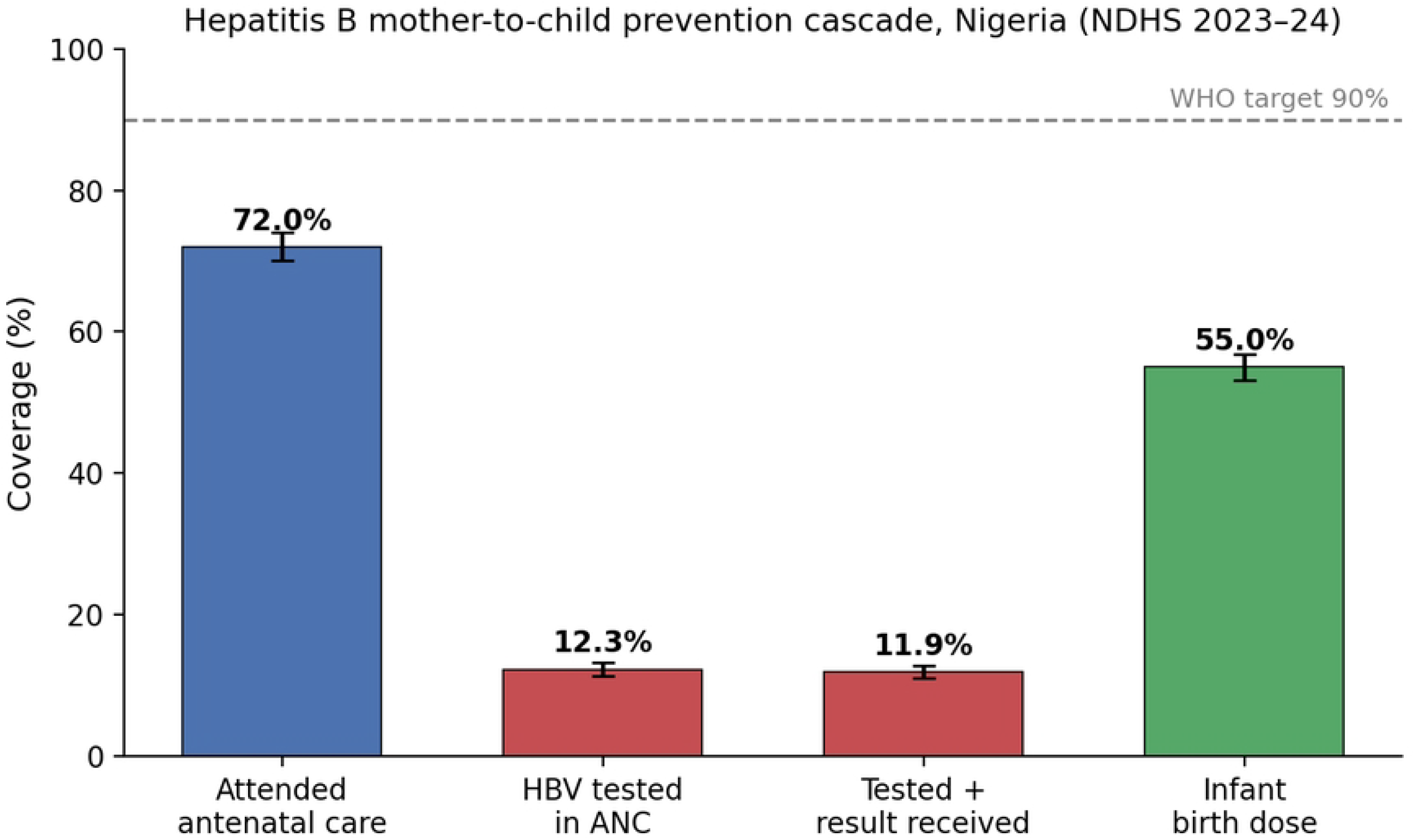
The hepatitis B mother-to-child prevention cascade, Nigeria (NDHS 2023–24). Weighted coverage of each cascade step among recent mothers, with 95% confidence intervals; dashed line marks the WHO triple-elimination target of 90%. Antenatal attendance (blue) is high, hepatitis B testing (red) collapses on that same platform, and the infant birth dose (green) sits at a separate level, reflecting its universal, non-targeted delivery.

### Who the cascade reaches

Both steps were steeply patterned by wealth, education, and remoteness, and the gradients ran in parallel (Table 2). Antenatal testing rose from 2.6% (95% CI 1.8–3.4) in the poorest wealth quintile to 32.4% (29.8–35.0) in the richest, and from 3.7% among women with no education to 38.8% among those with higher education; it fell from 24.1% in the most accessible areas to 4.8% in the most remote. The birth dose traced the same contours: 33.5% to 87.8% across wealth, 33.1% to 89.0% across education, and 76.0% down to 35.0% from least to most remote. The regional pattern compounded these gradients: the North-West, the zone with Nigeria’s highest hepatitis B prevalence [2], and the neighbouring North-East recorded the lowest testing (7.7% and 8.3%) and, in the North-West, the lowest birth dose (36.1%), while the southern zones recorded 11.3–22.3% testing and 79.5–84.9% birth dose. The two gradients do not run in step: on the birth dose the split is clean, every northern zone below 59% and every southern zone above 79%, whereas on testing the South-East (11.3%) sat below North-Central (18.5%). Testing was scarce almost everywhere; it was the birth dose that divided north from south. The women least reached at every step were the poorest, least-educated, and most remote — and those living where the virus is most common.

**Table 2.**
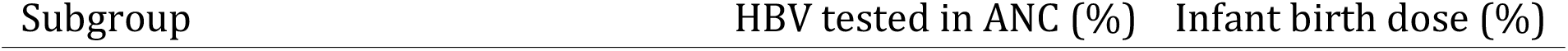
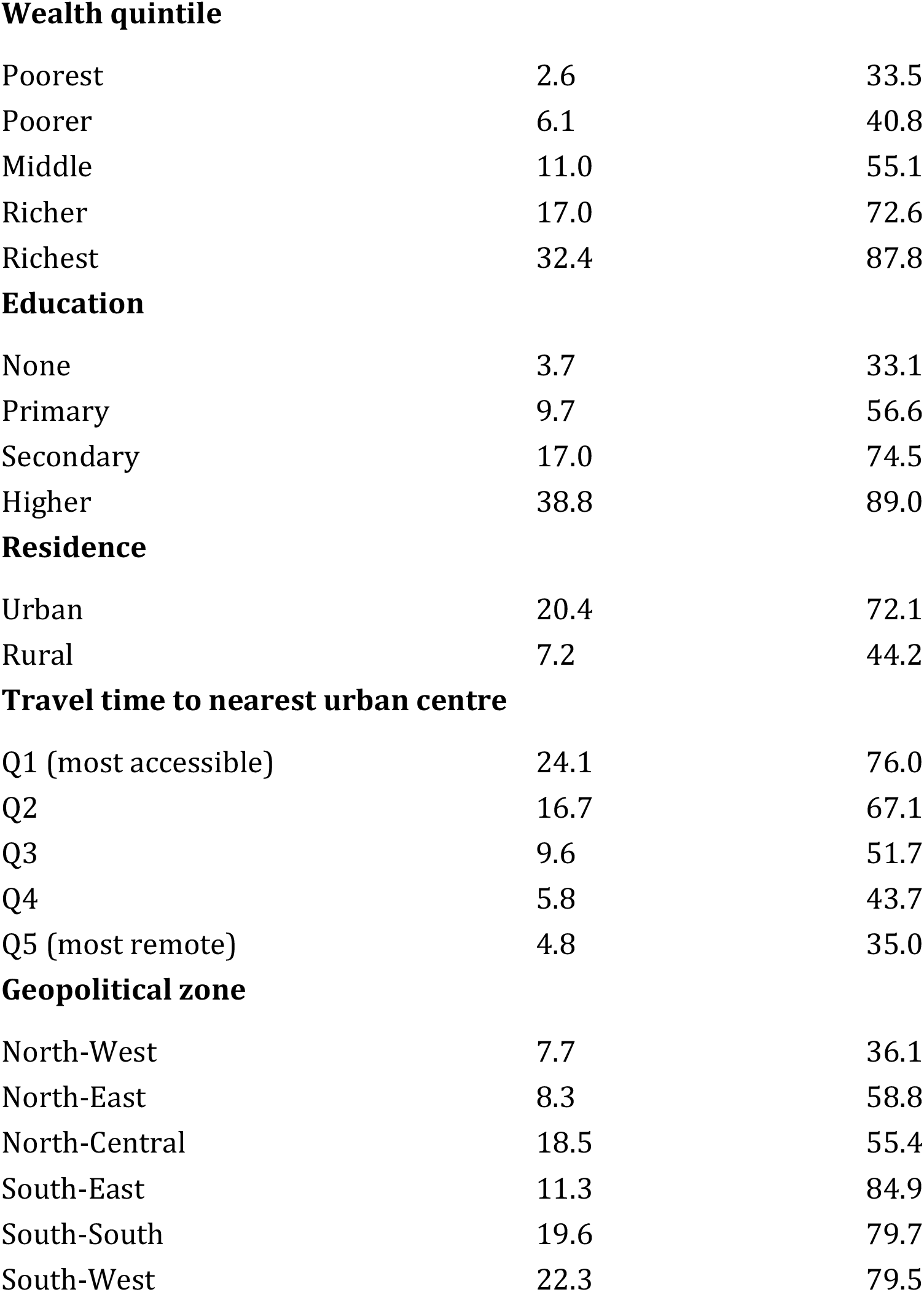
Antenatal hepatitis B testing and infant birth-dose coverage by socioeconomic and geographic subgroup (weighted %), NDHS 2023–24.

| Subgroup | HBV tested in ANC (%) | Infant birth dose (%) |
| --- | --- | --- |
| **Wealth quintile** |  |  |
| Poorest | 2.6 | 33.5 |
| Poorer | 6.1 | 40.8 |
| Middle | 11.0 | 55.1 |
| Richer | 17.0 | 72.6 |
| Richest | 32.4 | 87.8 |
| **Education** |  |  |
| None | 3.7 | 33.1 |
| Primary | 9.7 | 56.6 |
| Secondary | 17.0 | 74.5 |
| Higher | 38.8 | 89.0 |
| **Residence** |  |  |
| Urban | 20.4 | 72.1 |
| Rural | 7.2 | 44.2 |
| **Travel time to nearest urban centre** |  |  |
| Q1 (most accessible) | 24.1 | 76.0 |
| Q2 | 16.7 | 67.1 |
| Q3 | 9.6 | 51.7 |
| Q4 | 5.8 | 43.7 |
| Q5 (most remote) | 4.8 | 35.0 |
| **Geopolitical zone** |  |  |
| North-West | 7.7 | 36.1 |
| North-East | 8.3 | 58.8 |
| North-Central | 18.5 | 55.4 |
| South-East | 11.3 | 84.9 |
| South-South | 19.6 | 79.7 |
| South-West | 22.3 | 79.5 |

### Robustness

The findings held under alternative specifications. Antenatal testing coverage remained far below target even when “don’t know” responses were dropped from the denominator (20.3% versus 12.3%). In a supplementary analysis using card-documented vaccination, birth-dose coverage did not differ between infants of tested and untested mothers after accounting for their differing circumstances (adjusted prevalence ratio 1.065, 95% CI 0.996–1.137), consistent with a universal rather than risk-targeted birth dose. Place of delivery, by contrast, was strongly associated with the birth dose: infants born in a health facility were more likely to receive it (adjusted prevalence ratio 1.27, 95% CI 1.21–1.34), the largest association of any factor in the model. In the wider analysis that also counted maternal report, the residual association with maternal testing was slightly larger (adjusted prevalence ratio 1.09, 1.04–1.13) but fragile: an unmeasured confounder would need to be associated with both testing and the birth dose by a risk ratio of 1.39 to explain it away, and 1.26 to move the confidence bound to the null. Too few women reported a positive antenatal result for any assessment of whether they went on to receive antiviral prophylaxis.

## Discussion

### The main finding

This study followed recent Nigerian mothers from the antenatal clinic to their newborn’s first vaccination, and scored every step in between against the WHO triple-elimination targets. The platform needed to interrupt transmission is widely used and almost never used for the purpose. That coverage would fall short of those targets was expected; the size of the shortfall on testing was not. The country falls short by roughly 78 points on antenatal testing and 35 on the birth dose, and the shortfall is deepest in the North-West, the zone where hepatitis B is most prevalent [2], and in the neighbouring North-East. The prevention cascade is not so much leaking as largely unbuilt.

### Why testing is so rare on a platform most women reach

The barriers are practical and documented. Nigerian primary-care workers point to absent test kits, too few trained staff, and, repeatedly, the out-of-pocket cost of the test itself [13]. Testing is not routinely free, and where it is charged it competes with every other pregnancy expense. The cost barrier does not stop at the test: even the immunoglobulin that would protect an exposed newborn is priced beyond most families, and although 86% of pregnant women in one Nigerian study were willing to pay for it, the most they would offer averaged 23,178 naira (about US$63), far below its market price [14]. If the constraint were the women themselves, this is the platform on which testing should already be routine: seven in ten attend, and they attend for care. Contrary to that reading, the evidence from comparable settings places the constraint on the supply side. In four rural primary health centres in western Uganda, staffed by clinical officers, nurses and midwives rather than physicians, all 1,065 women offered hepatitis B screening at their first antenatal visit accepted it, and 47 of the 49 who tested positive went on to complete viral load testing [15]. Ugandan guidelines had recommended antenatal screening all along; what prevented it was stockouts of rapid tests and the absence of any pathway for managing a positive result. Where the test was supplied and the pathway built, uptake was complete. That study also marks the limit of what supply alone buys: viral load results took a median of 46 days to return, in pregnancies already at 22 weeks when screening took place. Layered onto this is a division of labour our data make visible: the birth dose is given by immunisation staff whose charge is the child, independent of any maternal test, so a mother’s hepatitis B status, if it is ever established, has no obvious route into her infant’s care. Two parallel systems result, one for vaccinating babies and one, barely functioning, for testing mothers, that do not meet. What decides whether an infant is vaccinated is therefore not what was learned about the mother but where she gave birth: facility delivery carried the largest association with the birth dose of anything we modelled, far larger than the residual for maternal testing. The incomplete birth dose is, on this reading, a problem of where labour happens rather than of what antenatal care discovers.

### What this adds

These findings extend what was known from smaller or single-step studies. A single-state Nigerian cohort had found birth-dose coverage unrelated to maternal hepatitis B status [3]; we show, nationally, that the more fundamental problem lies upstream — most mothers are never tested at all. Reviews had argued from programme descriptions that selective birth-dose vaccination cannot interrupt transmission without antenatal screening [4], and had called for evaluation of the full cascade in West Africa, where such evidence was absent [9]; we provide that national cascade. The clinical rationale is not in doubt: even a timely birth dose leaves a residual transmission risk concentrated in high-viral-load mothers, who can be identified and treated only if they are tested in pregnancy [7]. A country testing one in eight pregnant women cannot mount that risk-targeted prevention at all.

### Implications

The remedy lies in fuller use of the platform that already reaches most women. Integrating hepatitis B testing into routine antenatal care, ideally within a single multiplex test alongside HIV, syphilis, and malaria, is both feasible and cheap. A four-infection antenatal panel delivered from a single fingerstick averted 92 disability-adjusted life years per 1,000 pregnant women across Kenya, Rwanda, and Uganda, at US$5.76 to US$32.62 per DALY averted, and was cost-saving outright in Uganda [16]. Hepatitis B testing and treatment are broadly cost-effective even where prevalence is modest [17].

Making the test free at the point of care, supplying kits, and linking a positive result to the infant’s birth dose and to maternal antiviral prophylaxis would be the step most likely to convert antenatal contact already achieved into protection. That the cascade should track wealth and remoteness was expected; that it should be weakest precisely where the virus is commonest was not, and it is what turns a coverage problem into an equity one. The equity pattern sharpens the priority: because testing and the birth dose both collapse in the poorest, most remote, and highest-prevalence north, an untargeted national scale-up could widen the very gap it aims to close; delivery must be weighted toward those settings.

### Limitations

Several bounds apply. Testing, results, and the birth dose are self-reported; mothers may over-report each, so the coverage figures are best read as upper bounds and the true gaps as at least as wide, an interpretation reinforced by the birth dose falling further when restricted to card-documented receipt. Timeliness of the birth dose, on which protection depends, is not recorded, nor is whether immunoglobulin or maternal antivirals were given; we therefore describe coverage, not averted transmission. The management of high-risk mothers is therefore invisible here: the women who most needed prophylaxis are the ones this survey counts worst. Because the antenatal questions refer to the most recent birth, the cascade speaks to recent mothers rather than all women. And travel time is a modelled proxy for access, not a measured distance to a facility that offers testing. None of these would erase a shortfall of the magnitude observed.

### Conclusion

Nigeria already has, in its antenatal services, the reach required to interrupt mother-to-child transmission of hepatitis B, and to prevent the liver cancer that follows. What it lacks is the test on that platform and the completion of the dose behind it. Closing a preventable-cancer gap here is less a matter of building new contact than of using the contact already made, and of doing so first where the virus, and the disadvantage, run deepest.

## Data Availability

The data underlying this study are third-party data owned by The DHS Program and are available at https://dhsprogram.com/data/ following free registration and a brief description of the intended analysis. The authors received no special access privileges and confirm that other researchers can access the data in the same manner. The variables and coding rules needed to reproduce every reported estimate are provided in S1 Table.

## Acknowledgments

The authors thank The DHS Program and the National Population Commission of Nigeria for collecting and making available the 2023-24 Nigeria Demographic and Health Survey, and the women who gave their time to the survey.

## Supporting information

**S1 Table. Survey variables and coding rules**. The NDHS 2023-24 variables, skip patterns and coding decisions behind every cascade step, analytic population, covariate and sensitivity analysis reported in this paper.

**S2 Table. Full cascade estimates**. Numerators, denominators and weighted percentages for each cascade step and population, with 95% confidence intervals by Taylor-series linearisation, each cross-checked against a delete-one-primary-sampling-unit jackknife.

**S3 Table. Supplementary regression models**. Modified Poisson models for the infant birth dose, crude through fully adjusted, with the place-of-delivery association and E-values for unmeasured confounding.

**S4 Table. Sensitivity analyses**. Alternative coding of “don’t know” responses, card-documented birth dose only, an alternative geographic access measure, and the counts underlying each.

**S5 File. Analysis code**. Annotated R script `(verify_cascade.R)` that locates the survey files, reproduces every estimate reported in this article and its supporting tables, and prints a verification table comparing each value against the manuscript.

